# A model to predict SARS-CoV-2 infection based on the first three-month surveillance data in Brazil

**DOI:** 10.1101/2020.04.05.20047944

**Authors:** Fredi A. Diaz-Quijano, José Mário Nunes da Silva, Fabiana Ganem, Silvano Oliveira, Andrea L. Vesga-Varela, Julio Croda

## Abstract

**Background:** COVID-19 diagnosis is a critical problem, mainly due to the lack or delay in the test results. We aimed to obtain a model to predict SARS-CoV-2 infection in suspected patients reported to the Brazilian surveillance system.

**Methods:** We analyzed suspected patients reported to the National Surveillance System that corresponded to the following case definition: patients with respiratory symptoms and fever, who traveled to regions with local or community transmission or who had close contact with a suspected or confirmed case. Based on variables routinely collected, we obtained a multiple model using logistic regression. The area under the receiver operating characteristic curve (AUC) and accuracy indicators were used for validation.

**Results:** We described 1468 COVID-19 cases (confirmed by RT-PCR) and 4271 patients with other illnesses. With a data subset, including 80% of patients from Sao Paulo (SP) and Rio Janeiro (RJ), we obtained a function which reached an AUC of 95.54% (95% CI: 94.41% - 96.67%) for the diagnosis of COVID-19 and accuracy of 90.1% (sensitivity 87.62% and specificity 92.02%). In a validation dataset including the other 20% of patients from SP and RJ, this model exhibited an AUC of 95.01% (92.51% – 97.5%) and accuracy of 89.47% (sensitivity 87.32% and specificity 91.36%).

**Conclusion:** We obtained a model suitable for the clinical diagnosis of COVID-19 based on routinely collected surveillance data. Applications of this tool include early identification for specific treatment and isolation, rational use of laboratory tests, and input for modeling epidemiological trends.

## INTRODUCTION

The pandemic caused by the novel coronavirus, Sars-Cov-2, challenges the capabilities of health care services, especially in low- and middle-income countries [1]. A major issue is to meet the diagnostic requirements of the suspected cases reported to the surveillance system [2]. The proportion of suspected cases being tested in each country is not systematically presented in most of the epidemiological reports [3–5]. However, with the increasing number of new suspected cases of the disease (COVID-19) worldwide, the diagnosis has become a growing problem, mainly due to the lack or delay in the test results [6,7].

Clinical manifestations of COVID-19 are unspecific and include respiratory symptoms, fever, cough, dyspnea, and viral pneumonia [8–10]. Polymerase chain reaction by real-time reverse transcriptase (RT-PCR) is considered the gold standard for the diagnosis of SARS-CoV-2 infection. However, its limited availability and the strict laboratory requirements delay diagnosis, which represents an unprecedented challenge to control transmission and provide timely health care [11,12].

The incorporation of predictive diagnostic models based on surveillance data could help identify patients who could need specific treatment and early isolation. Consequently, we aimed to describe the profile of COVID-19 patients and to obtain a multiple model to predict the diagnosis among suspected cases reported in Brazil, based on data routinely collected by the surveillance system.

## METHODS

### Study design and population

This observational study corresponded to a developing and evaluation of diagnostic technologies, nested in surveillance data obtained by the Brazilian Ministry of Health. We studied the reported cases, which corresponded to the following case definition: patients with respiratory symptoms and fever, who had traveled to regions with community or local transmission or who had close contact with a suspected or confirmed case. We did not establish restrictions based on age or underlying conditions for this study. Records with inconsistent or illogical data were excluded.

### Procedures

We included the patients reported between 11/01/2020 and 25/03/2020. All data were collected from the national surveillance information form, created on RedCap® platform, which included demographic, temporal, and easy to obtain clinical information such as symptoms, signs, comorbidities, travel history, and contact information. Another variable considered was the time since notification of the first case that was subsequently confirmed in the corresponding Federal Unit (FU). In the FU without notification and for its first confirmed case, this variable was zero.

During the period of data collection analyzed, the ministry’s recommendation was to test all suspected cases, according to the definition presented above [13]. SARS-CoV-2 infection was considered confirmed only by Real-time reverse-transcriptase polymerase chain reaction RT-PCR testing, following the WHO and CDC protocol result for pharyngeal swab specimens [12]. Because the study population refers to symptomatic cases, in this paper we used the terms SARS-CoV-2 infection and COVID-19 interchangeably.

The reference definition of COVID-19 was a reported case with a RNA test positive. Because the target population was the suspected cases identified through the surveillance system, the COVID-19 group was compared with the reported cases with RNA test negative (henceforth named other illness [OI] group).

### Data analysis

Demographic and clinical information was entered in an electronic database and then analyzed using Excel and STATA (version 15.0, Stata Corp LP, College Station, TX, USA). Data analysis included a description of the manifestations of the disease, according to etiology (COVID-19 vs. OI). During the univariable analysis, we sought the most functional form of the available variables. This included the evaluation of composite variables for categorical predictors and the evaluation of the linear relationship between quantitative variables and the frequency of the outcome.

Age showed a biological gradient with COVID-19; therefore, a simple imputation was made for cases with missing values of this variable, considering the frequency of the diagnosis. Thus, the value of 37.38 years was calculated to impute unregistered age (in 1.4% of patients with etiological diagnosis). This value corresponded to the expected age (*A*_*e*_), considering the COVID-19 odds in the missing group (*Odds*_*m*_) and the coefficient *(β*_*age*_) of a simple logistic regression of this outcome on age in the other patients. For this imputation, we took as reference values the odds (*Odds*_*r*_) and mean age (*A*_*r*_) of the decile with the odds closest to that of the missing-age group (sixth decile, aged between 34.3 and 38.1, mean: 36.28 years old). Explicitly,

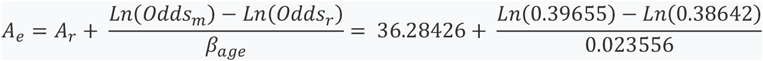

The information from São Paulo (SP) and Rio de Janeiro (RJ) was used to obtain and validate the predictive model. This choice was because these were the FUs with the largest number of confirmed cases and the earliest establishment of the surveillance system. Thus, we used a subset of 80% of randomly selected patients from SP and RJ (modeling dataset) to specify the multiple model. We selected the covariates by a non-automatic stepwise procedure using logistic regression. Age and days after notification of the first confirmed case (DNFCC) were used to create interaction terms with each of the other independent predictors. During modeling, a p-value of 0.15 was considered as a criterium to enter the variable and 0.20 to exclude it. After evaluating all the variables, exclusions were made until obtaining a model including only covariates with p <0.10. To obtain a final predictive function we integrated the multiple model and variables that perfectly predicted the outcome.

The predicted values were used to estimate the area under the ROC curve (AUC). We interpreted the AUC as an indicator of goodness of fit such that values between 0.9 and 0.99 are excellent, 0.8 – 0.89 good, 0.7 – 0.79 acceptable, and 0.51 – 0.69 are poor [14]. We also plotted the observed COVID-19 proportion by decile of predictions to illustrate the calibration of the model. Next, the model was applied to the 20% of patients from SP and RJ who were not included in the modeling dataset (validation dataset). We chose that sample distribution (80% and 20%) looking to have more than 100 events in the validation dataset [15]. Moreover, we applied the model to patients from FUs other than SP/RJ to evaluate the applicability in a very different scenario. We also calculated the overall accuracy to classify events of a predicted probability of ≥0.5 [16].

We presented some cut-off points of the predicted value based on optimized accuracy indicators (in SP/RJ patients). These cut-offs included: the preset predicted value of 0.5; the highest value with a sensitivity >95%; the lowest with specificity >95%; the value with the highest overall accuracy; and the value with the best balance between sensitivity and specificity (based on the product thereof). Accuracy indicators of these selected cutoffs were described for both the SP/RJ patients (modeling + validation dataset) and those from the other FUs.

Finally, by applying the sum of the predicted values and by using the chosen cut-off points, we calculated the probable number of COVID-19 cases in the total reported patients and among those who were reported as being hospitalized.

## RESULTS

Until March 25, 2020, the surveillance system had received 67,344 records of suspected cases, including 5674 with registered hospitalization. Of the total, 165 records were excluded because of inconsistent data. Overall, 5739 were tested by RT-PCR, of which 1468 were positive and 4271 negative.

COVID-19 cases were older and more frequently men compared with OI patients (Table 1). COVID-19 patients were reported in median 16 days after the first confirmed case, which was significantly later than OI patients were (median of seven days). Both age and time from the first confirmed case exhibited a gradient for the COVID-19 frequency (Figures 1 and 2).

**Table 1.** Comparison of COVID-19 patients and other illnesses (OI) reported to the Brazilian surveillance system.

| Variable | Total<br>(n= 5739) | COVID-19<br>(n=1468) | OI<br>(n=4271) | p-value <sup>a</sup> |
| --- | --- | --- | --- | --- |
| Age (years) - median (IQR) <sup>b</sup> | 35.4 (26.5 – 48.2) | 39.6 (31 – 53.5) | 33.7 (25.1 – 46) | <0.001 <sup>c</sup> |
| Sex – Female | 3037 (52.9%) | 662 (45.1%) | 2375 (55.6%) | <0.001 |
| Male | 2600 (45.3%) | 776 (52.9%) | 1824 (42.7%) |  |
| Unregistered | 102 (1.8%) | 30 (2%) | 73 (1.7%) |  |
| DARFCC <sup>d</sup> — Median (IQR) | 9 (2 – 16) | 16 (9 – 20) | 7 (1 – 13) | <0.001 <sup>c</sup> |
| **Symptoms** |  |  |  |  |
| Fever | 4368 (76.1%) | 982 (66.9%) | 3386 (79.3%) | <0.001 |
| Cough | 4577 (79.8%) | 1040 (70.8%) | 3537 (82.8%) | <0.001 |
| Sore throat | 2816 (49.1%) | 483 (32.9%) | 2333 (54.6%) | <0.001 |
| Breathing difficulty | 1353 (23.6%) | 231 (15.7%) | 1122 (26.3%) | <0.001 |
| Myalgia or arthralgia | 1431 (24.9%) | 450 (30.7%) | 981 (23%) | <0.001 |
| Diarrhea | 599 (10.4%) | 117 (8%) | 482 (11.3%) | <0.001 |
| Nausea or vomiting | 429 (7.5%) | 74 (5%) | 355 (8.3%) | <0.001 |
| Headache | 1948 (33.9%) | 433 (29.5%) | 1515 (35.5%) | <0.001 |
| Coryza | 2797 (48.7%) | 495 (33.7%) | 2302 (53.9%) | <0.001 |
| Irritability or confusion | 73 (1.3%) | 14 (1%) | 59 (1.4%) | 0.21 |
| Adynamia or weakness | 924 (16.1%) | 224 (15.3%) | 700 (16.4%) | 0.31 |
| Sputum | 341 (5.9%) | 44 (3%) | 297 (7%) | <0.001 |
| Chills | 608 (10.6%) | 152 (10.4%) | 456 (10.7%) | 0.73 |
| Nasal congestion | 1045 (18.2%) | 228 (15.5%) | 817 (19.1%) | 0.002 |
| Conjunctival congestion | 113 (2%) | 17 (1.2%) | 96 (2.2%) | 0.01 |
| Difficulty swallowing | 137 (2.4%) | 18 (1.2%) | 119 (2.8%) | <0.001 |
| Red spots on the body | 32 (0.6%) | 3 (0.2%) | 29 (0.7%) | 0.04 <sup>e</sup> |
| Enlarged lymph nodes | 45 (0.8%) | 8 (0.5%) | 37 (0.9%) | 0.30 <sup>e</sup> |
| Nasal wing beat | 25 (0.4%) | 2 (0.1%) | 23 (0.5%) | 0.06 <sup>e</sup> |
| Oxygen saturation <95 | 122 (2.1%) | 37 (2.5%) | 85 (2%) | 0.22 |
| Signs of cyanosis | 17 (0.3%) | 1 (0.1%) | 16 (0.4%) | 0.09 <sup>e</sup> |
| Intercostal circulation | 17 (0.3%) | 3 (0.2%) | 14 (0.3%) | 0.59 <sup>e</sup> |
| Dyspnoea | 466 (8.1%) | 111 (7.6%) | 355 (8.3%) | 0.36 |
| Other symptoms | 683 (11.9%) | 151 (10.3%) | 532 (12.5%) | 0.03 <sup>e</sup> |
| **Signs** |  |  |  |  |
| Fever | 1268 (22.1%) | 267 (18.2%) | 1001 (23.4%) | <0.001 |
| Exudate pharyngeal | 283 (4.9%) | 42 (2.9%) | 241 (5.6%) | <0.001 |
| Convulsion | 4 (0.1%) | 1 (0.1%) | 3 (0.1%) | 1 <sup>e</sup> |
| Conjunctivitis | 70 (1.2%) | 10 (0.7%) | 60 (1.4%) | 0.03 <sup>e</sup> |
| Coma | 3 (0.1%) | 3 (0.2%) | 0 (0%) | 0.02 <sup>e</sup> |
| Dyspnoea or tachypnea | 518 (9%) | 90 (6.1%) | 428 (10%) | <0.001 |
| Alteration detected by pulmonary auscultation | 237 (4.1%) | 42 (2.9%) | 195 (4.6%) | 0.005 |
| Radiological alteration | 186 (3.2%) | 45 (3.1%) | 141 (3.3%) | 0.66 |
| Other signs | 896 (15.6%) | 147 (10%) | 749 (17.5%) | <0.001 |

**Table 1. Comparison of COVID-19 patients and other illnesses (OI) reported to the Brazilian surveillance system (continued).**
| Variable | Total<br>(n= 5739) | COVID-19<br>(n=1468) | OI<br>(n=4271) | p-value |
| --- | --- | --- | --- | --- |
| **Clinical history** |  |  |  |  |
| Cardiovascular disease<br>(including hypertension) | 475 (8.3%) | 116 (7.9%) | 359 (8.4%) | 0.55 |
| Diabetes | 195 (3.4%) | 41 (2.8%) | 154 (3.6%) | 0.14 |
| Liver disease | 16 (0.3%) | 0 (0%) | 16 (0.4%) | 0.02 <sup>e</sup> |
| Chronic neurological or<br>neuromuscular disease | 32 (0.6%) | 3 (0.2%) | 29 (0.7%) | 0.04 <sup>e</sup> |
| Immunodeficiency | 50 (0.9%) | 11 (0.7%) | 39 (0.9%) | 0.56 |
| HIV | 23 (0.4%) | 6 (0.4%) | 17 (0.4%) | 1 <sup>e</sup> |
| Renal disease | 29 (0.5%) | 4 (0.3%) | 25 (0.6%) | 0.20 <sup>e</sup> |
| Chronic pulmonary disease | 196 (3.4%) | 34 (2.3%) | 162 (3.8%) | 0.007 |
| Neoplasia | 57 (1%) | 16 (1.1%) | 41 (1%) | 0.66 |
| Claim not to have had contact<br>with a suspect case | 203 (3.5%) | 0 | 203 (4.8%) | <0.001 |
| Trip outside Brazil up to 14 days<br>before the onset of symptoms? |  |  |  |  |
| Yes | 3319 (57.8%) | 517 (35.2%) | 2802(65.6%) | <0.001 |
| Not | 2094 (36.5%) | 749 (51%) | 1345(31.5%) |  |
| Don't know or missing | 326 (5.7%) | 202 (13.8%) | 124 (2.9%) |  |

<sup>a</sup> Unless otherwise specified, p-values were obtained using the chi-square test.
<sup>b</sup> n=1,445 vs 4,213.
<sup>c</sup> Mann-Whitney test
<sup>d</sup> DARFCC: Days after the reporting of the first confirmed case.
<sup>e</sup> Fisher's exact test

**Figure 1.**
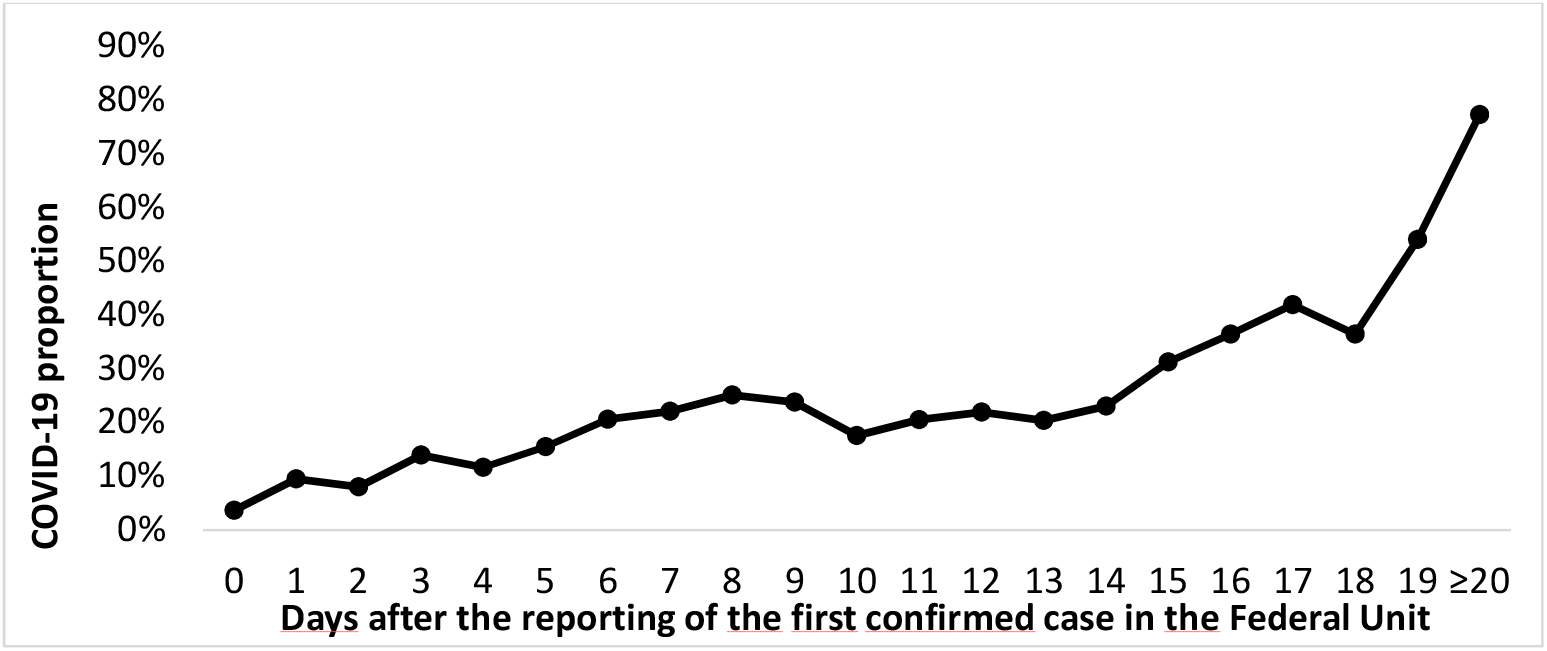
COVID-19 proportion among suspected cases according to time after the reporting of the first confirmed case.

**Figure 2.**
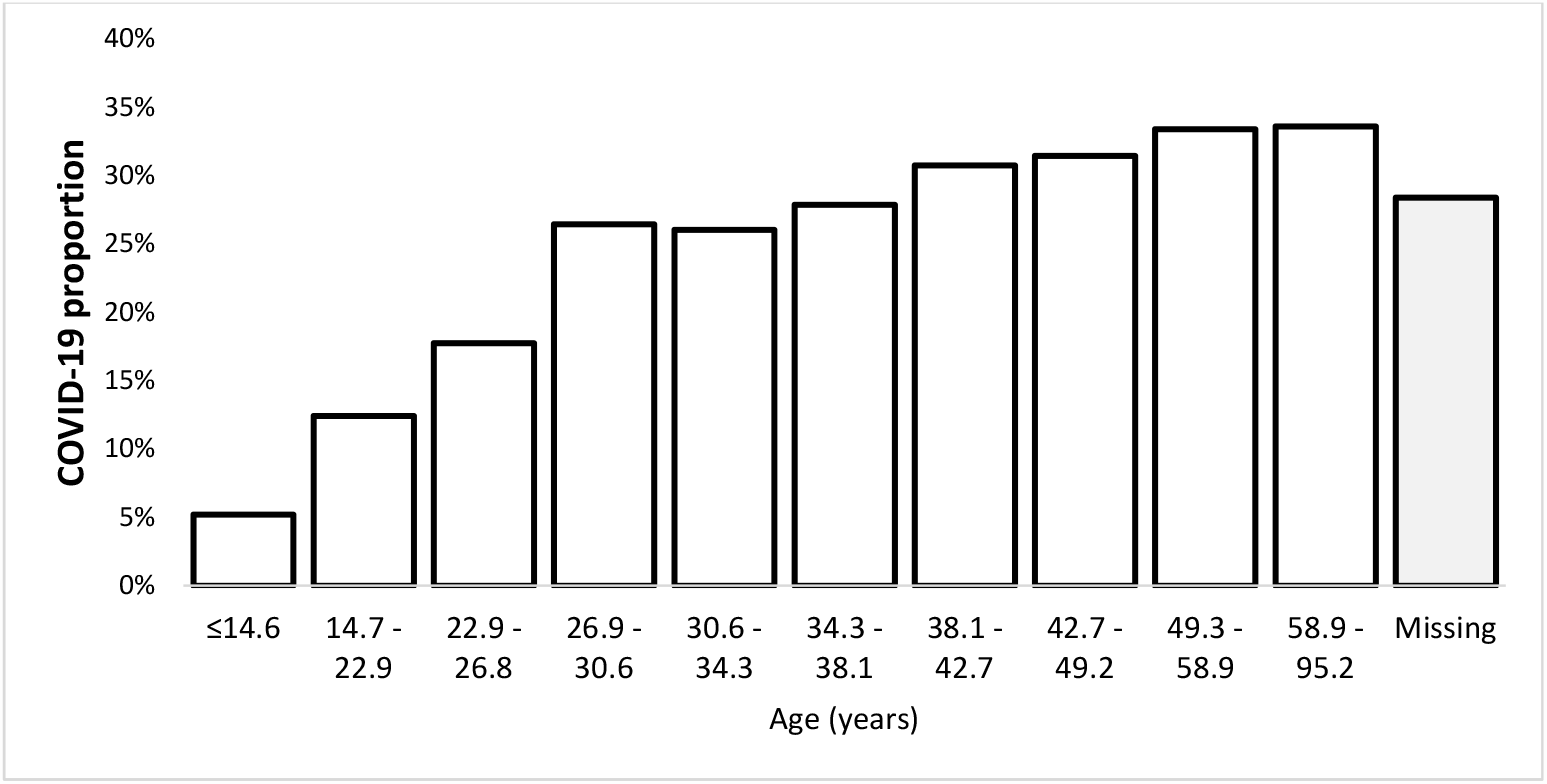
COVID-19 proportion among suspected patients according to age.

Most of the clinical manifestations were more frequent in OI patients than COVID-19 in the univariable analysis. Only the myalgia or arthralgia variable was significantly more frequent in COVID-19 than OI (30.7% vs. 23%, p<0.001). No COVID-19 infections were observed among patients with liver disease or among those that claimed not to have been in contact with a suspected case. On the other hand, COVID-19 patients less frequently referred to making a trip outside Brazil in the last 14 days (Table 1).

### Multiple model

The states of SP and RJ jointly had 683 confirmed COVID-19 cases and 864 with OI, of which 541 and 702 were selected to the modeling dataset, respectively. During the modeling, patients with liver disease (n = 4) and those who reported not having had contact with a suspected case (n = 69) were not considered, as these categories perfectly predicted absence of COVID-19 and were significantly more frequent in the OI group (Table 1).

We obtained a model integrating 15 covariates, including age, days from notification of the first confirmed case (DNFCC) in the corresponding FU, eight variables about clinical manifestations, two on comorbidities, trip history, and two interaction terms (Table 2). The AUC of this multiple model was estimated at 95.36% (95% CI: 94.2 – 96.52%) with an accuracy of 89.5%.

**Table 2.** Predictive model for COVID-19 diagnoses among reported patients.

|  |  |  |
| --- | --- | --- |
| **Variable** | **OR (95% CI)** | **p value** |
| Age (in years) | 1 (0.98 - 1.02) | 0.86 |
| DARFCC <sup>a</sup> | 1.46 (1.39 - 1.54) | <0.001 |
| Fever (symptom) | 0.17 (0.05 - 0.56) | 0.003 |
| Age * Fever <sup>b</sup> | 1.03 (1 - 1.06) | 0.03 |
| Cough (symptom) | 0.47 (0.29 - 0.74) | 0.001 |
| Sore throat (symptom) | 0.47 (0.31 - 0.7) | <0.001 |
| Diarrhea (symptom) | 0.1 (0.01 - 0.87) | 0.04 |
| Age * Diarrhea <sup>b</sup> | 1.07 (1.02 - 1.13) | 0.01 |
| Coryza (symptom) | 0.45 (0.3 - 0.67) | <0.001 |
| Chills (symptom) | 1.85 (0.98 - 3.51) | 0.06 |
| Pulmonary manifestation <sup>c</sup> | 0.43 (0.26 - 0.71) | 0.001 |
| Other signs | 0.46 (0.25 - 0.86) | 0.02 |
| HIV | 19.8 (0.85 - 462.81) | 0.06 |
| Kidney disease | 0.06 (0 - 1.06) | 0.06 |
| Trip outside Brazil up to 14 days before the onset of symptoms? | . | . |
| Not | 3.11 (2 - 4.82) | <0.001 |
| Don't know or missing | 3.02 (1.06 - 8.58) | 0.04 |
| Intercept | 0.02 (0.01 - 0.07) | <0.001 |

<sup>a</sup> Days after the reporting of the first confirmed case.
<sup>b</sup> Interaction term defined by the multiplication of variables.
<sup>c</sup> Composite variable defined as any breathing difficulty, dyspnea (symptom or sign), tachypnea, or pulmonary alteration detected by auscultation.

To obtain the final function, patients with a history of liver disease and those who denied having had any contact with a suspected case were considered with a predicted value equal to zero; otherwise, the predicted value was calculated by applying the model described in table 2. With this predictive function, the area was 95.54% (95% CI: 94.41% – 96.67%) for the diagnosis of COVID-19 in the modeling dataset and 95.01% (92.51% – 97.5%) in the validation dataset (Figure 3). Accuracy in these datasets was 90.1% (sensitivity 87.62% and specificity 92.02%) and 89.47% (sensitivity 87.32% and specificity 91.36%), respectively. Furthermore, the calibration plots suggested reliable predictive performance in the validation group, similar to displayed in the modeling dataset (Figure 4).

**Figure 3.**
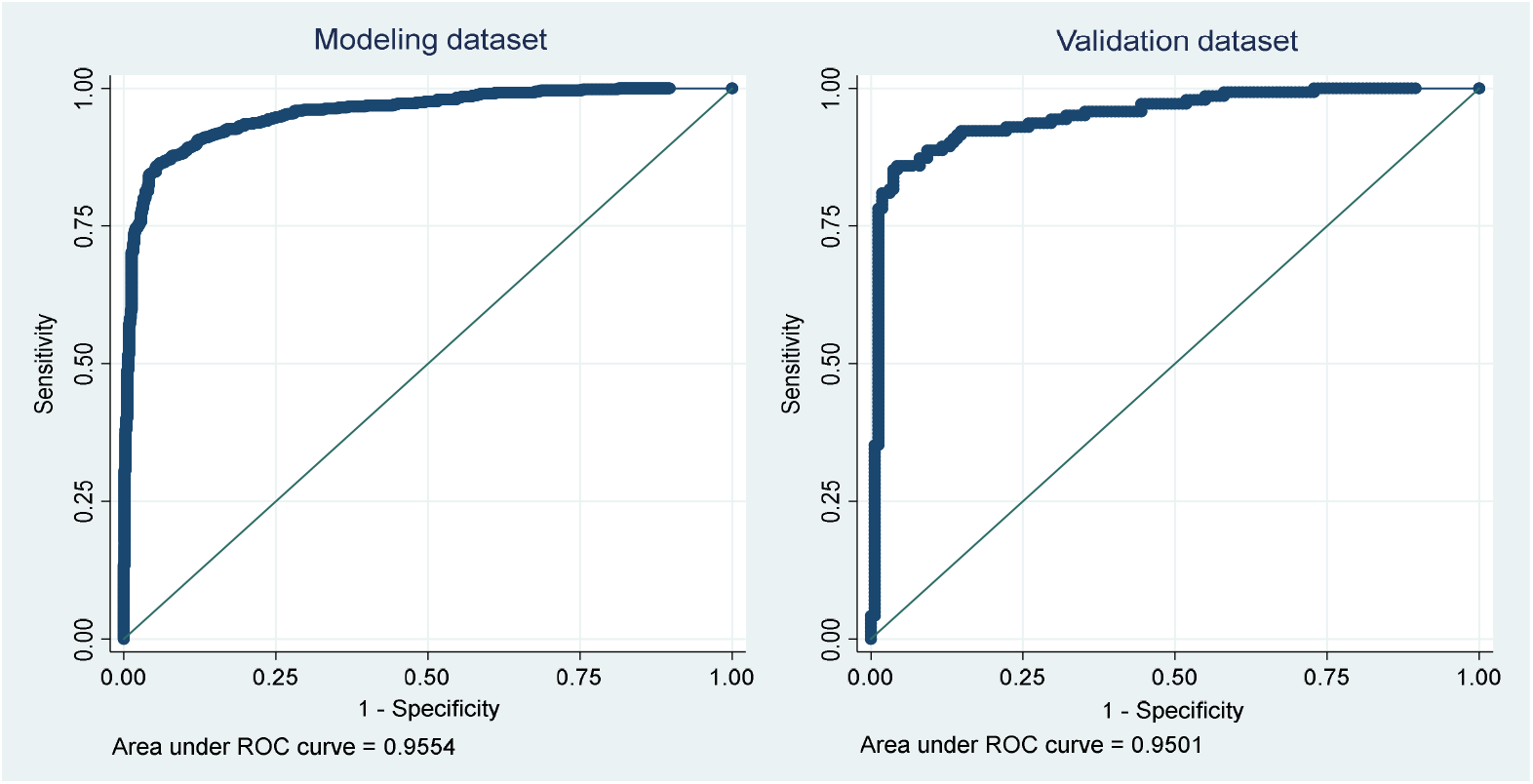
Area under the ROC curve in the modeling and validation datasets.

**Figure 4.**
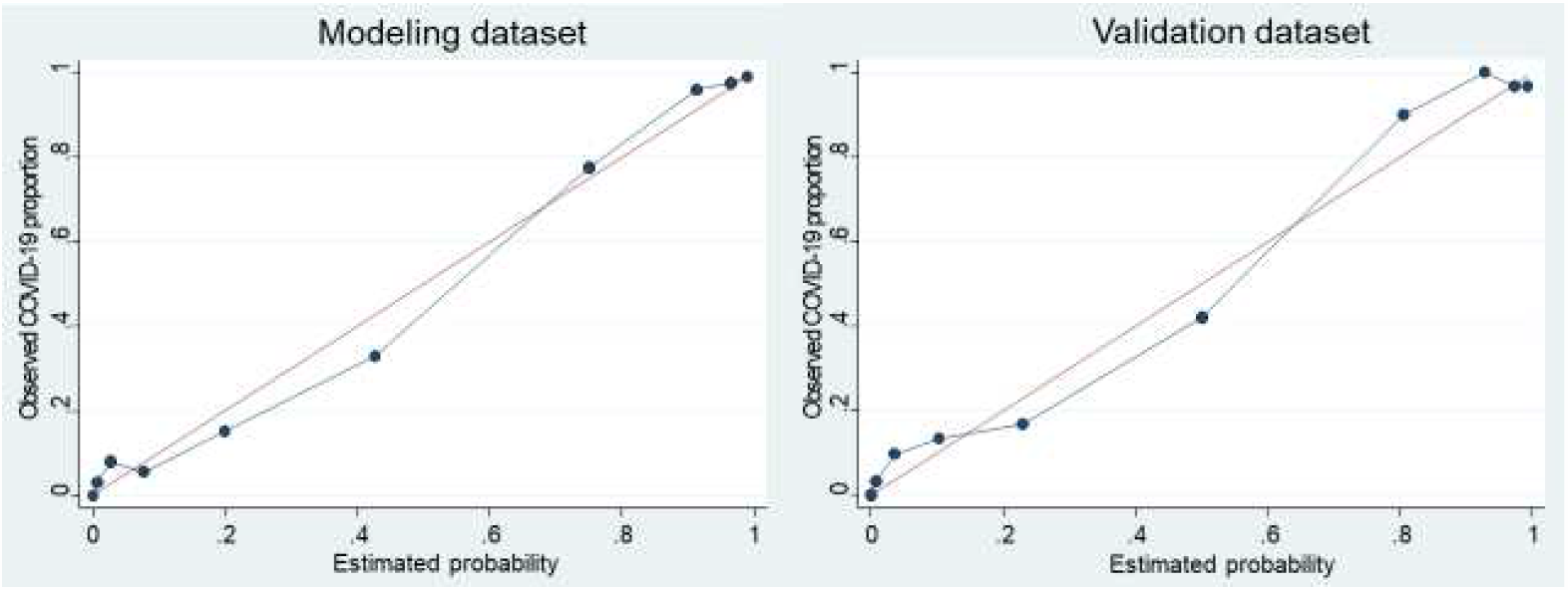
Calibration plots in the modeling and validation datasets.

When this function was applied in patients from the other FUs, which included 785 cases of COVID-19 compared with 3407 with other diseases, the ROC area was 73.16% (95% CI: 71.35 – 74.96%), and the accuracy was 73.43% (sensitivity 46.37% and specificity 79.66%). In table 3, we described the diagnostic accuracy indicators of selected predicted-value cutoffs in both the SP/RJ and the other FU groups. Considering the sum of predicted values as well as the different cutoffs, the number of COVID-19 cases among reported patients (adding together confirmed and predicted by our model) would be between 22826 and 25190 in SP/RJ, and between 22704 and 28837 in other FUs (Table 4). Of them, between 2050 and 2196 were hospitalized in SP or RJ, and between 1657 and 2196 in the other FUs. All the calculations suggested that more than 95% of COVID-19 cases have not been confirmed.

**Table 3.** Diagnostic accuracy indicators of the selected predicted-value cutoff.

|  |  |  |  |  |  |  |  |  |  |  |  |
| --- | --- | --- | --- | --- | --- | --- | --- | --- | --- | --- | --- |
| **Criterion and value of cutoffs** |  | **SP and RJ group** |  |  |  |  | **Other Federal Units group** |  |  |  |  |
| **Primary criterium** | **Cutoff** | **Sensitivity** | **Specificity** | **PPV** | **NPV** | **Accuracy** | **Sensitivity** | **Specificity** | **PPV** | **NPV** | **Accuracy** |
| Sensitivity $\geq 95\%$ | $\geq 0.1719$ | 95.0% | 73.0% | 73.6% | 94.9% | 82.7% | 63.6% | 66.8% | 30.6% | 88.8% | 66.2% |
| Prefixed | $\geq 0.5$ | 87.6% | 91.9% | 89.5% | 90.3% | 90.0% | 46.4% | 79.7% | 34.4% | 86.6% | 73.4% |
| Best balance (Sen*Spec)* | $\geq 0.5835$ | 85.9% | 94.7% | 92.7% | 89.5% | 90.8% | 44.3% | 82.1% | 36.4% | 86.5% | 75.0% |
| Specificity $\geq 95\%^*$ | $\geq 0.5956$ | 85.5% | 95.0% | 93.1% | 89.2% | 90.8% | 43.8% | 82.9% | 37.1% | 86.5% | 75.6% |

\* These cutoffs exhibited the highest accuracy in the SP/RJ group.

**Table 4.** Predicted cases and under-confirmation estimates of COVID-19 among suspected patients reported in Brazil, according to criteria based on the clinical predictive model.

| Primary criterium | All reported in SP and RJ |  |  | Hospitalized in SP and RJ |  |  | All reported in the other Federal Units |  |  | Hospitalized in the other Federal Units |  |  |
| --- | --- | --- | --- | --- | --- | --- | --- | --- | --- | --- | --- | --- |
|  | Additional predicted | Predicted + confirmed | Under-confirmation | Additional predicted | Predicted + confirmed | Under-confirmation | Additional predicted | Predicted + confirmed | Under-confirmation | Additional predicted | Predicted + confirmed | Under-confirmation |
| Sum of predicted | 22143 | 22826 | 97.0% | 1977 | 2050 | 96.4% | 22374 | 23159 | 96.6% | 1795 | 1869 | 96.0% |
| Sensitivity $\geq 95\%$ | 24507 | 25190 | 97.3% | 2123 | 2196 | 96.7% | 28052 | 28837 | 97.3% | 2196 | 2270 | 96.7% |
| Prefixed | 23303 | 23986 | 97.2% | 2029 | 2102 | 96.5% | 23298 | 24083 | 96.7% | 1770 | 1844 | 96.0% |
| Best balance (Sen*Spec) | 22852 | 23535 | 97.1% | 1996 | 2069 | 96.5% | 22089 | 22874 | 96.6% | 1673 | 1747 | 95.8% |
| Specificity $\geq 95\%$ | 22781 | 23464 | 97.1% | 1994 | 2067 | 96.5% | 21919 | 22704 | 96.5% | 1657 | 1731 | 95.7% |

## DISCUSSION

The growing number of cases suspected of COVID-19 is alarming [3]. Moreover, we observed that only a small proportion of the cases have a laboratory study. Therefore, most cases are being left with an uncertain diagnosis, which limits establishing specific measures and estimating the burden of the disease. In this study, we identified a set of variables that may help differentiate COVID-19 cases from other diseases. The model obtained exhibited an excellent AUC in the SP/RJ dataset comparable to more complex tools, including imaging and laboratory tests [11,17–19]. This is impressive, considering that it is based solely on variables collected by the surveillance system.

An essential caveat in these models is that the predictors should not be interpreted individually. However, some associations are consistent with what is known about this coronavirus. For example, age was directly associated with the diagnosis, which could be explained by the increased pathogenicity in older people. Therefore, an overrepresentation of the elderly is expected among the confirmed patients.

Another interesting finding is the relationship between the time since the notification of the first confirmed case and the probability of COVID-19. This association indicates the importance of contextualizing according to the timing of the epidemic. Furthermore, this demonstrates that these models should be continuously updated and adapted to the epidemiological situation.

Most of the clinical manifestations included in the model were negatively associated with the SARS-CoV-2 infection. It does not mean that they cannot be presented by patients with COVID-19, but that they were more frequent in other diseases. This finding highlights why the circulation of other infectious agents could be a determinant of the predictors’ discriminatory capacity, as has already been suggested for other conditions [20].

Moreover, it is expected that variables determining the notification (e.g., respiratory symptoms and international travel) and, therefore, inclusion in the study, tend to be negatively associated with the outcome due to collider-like phenomena [21]. For this reason, both causal inference interpretation, and extrapolation to the general population of the associations would be biased. Consequently, our model must be considered only for diagnostic prediction in the specific group of reported suspected patients.

The claim not to have had contact with an exposed case perfectly predicted the absence of COVID-19. This finding should be interpreted with caution because it is very likely that as the epidemic progresses, this variable could lose discrimination capacity once the prevalence of infectious hosts, including those undetectable, increases in the community.

Regarding external application, we observed that the model had a considerably lower AUC in FUs other than SP and RJ. This difference could occur due to the epidemiological context variability. For example, regional differences in the prevalence of respiratory pathogens other than SARS-CoV-2 can reduce the specificity of the clinical predictors. Moreover, the chronology of the COVID-19 epidemic itself (which started later in most of UFs other than in SP and RJ [22]) could affect its recognition as a public health priority and, therefore, the implementation and acceptability of its surveillance [23]. These factors could introduce heterogeneity of both the clinical profile of reported cases and the data recording quality, compromising the generalizability of the model performance. Despite this, the AUC in these other FUs can be considered acceptable, and although lower, the model proposed could also help guide the preliminary diagnosis in scenarios different than those obtained.

We take some steps to avoid biases frequently identified in models for diagnosis of COVID-19 [24]. For example, we did not exclude patients based on manifestations or evidence of other infections. In this way, our model could be applied to all reported to the surveillance systems during the emergency of COVID-19. In addition, the reference standard test was the same for all patients (RT-PCR), and no predictor was part of the outcome definition. Moreover, quantitative predictors were modeled without dichotomized them, allowing us to consider the continuous gradients of association with the outcome.

Applications of the proposed model include early case identification for specific treatment and isolation, as well as the rational use of laboratory tests. Furthermore, this model may predict the number of both total cases and hospitalizations attributed to this infection based on the surveillance data. This application is relevant because one of the challenges that this pandemic represents is the organization of healthcare resources. In this way, our results may help to model and forecast the availability of funds for patient care.

## Conclusions

This study obtained and validated a model function suitable for the clinical diagnosis of COVID-19 during the early stage of the Brazilian epidemic. This tool was entirely based on data routinely collected. Therefore, it may help early identification and treatment of patients, establish preventive measures, and improve the accuracy of epidemiological surveillance of this disease.

## Data Availability

Data subject to third party restrictions.

## Authors’ contributions

FADQ conceived the study, participated in its design and coordination, conducted the data analysis, and prepared the first draft of the manuscript. JMNS and ALVV helped plan the study, review of the literature, and variable codification. FG and SO worked on the collection and organization of the database. JC contributed in planning the study, database organization, and insights to the analysis process.

All authors provided relevant input for the writing, conducted reviews as well as read and approved the final manuscript. Thus, each author participated sufficiently in the work to take public responsibility for appropriate portions of the content and, therefore, agreed to be accountable for all aspects in ensuring that questions related to the accuracy or integrity of any part of the work are appropriately investigated and resolved.

## Acknowledgments

The authors thank Prof. Alexandre Dias Porto Chiavegatto Filho, director of the Big Data and Predictive Health Analysis Laboratory (Laboratório de Big Data e Análise Preditiva em Saúde - Labdaps) from FSP/USP, for his recommendations about the analysis plan.

## Ethical approval

This study followed Brazilian and International legislation for conducting human research. This research project was approved by the National Research Ethics Committee (Comissão Nacional de Ética em Pesquisa, CONEP) in Brazil, Register number (CAAE): 11946619.5.0000.5421.

